# Association of hypoxic-ischemic injury with MRI-derived glymphatic function markers in neonates: analysis of perivascular space volume and diffusion-tensor imaging

**DOI:** 10.1101/2025.03.12.25323818

**Authors:** Arum Choi, Dayeon Bak, Jimin Kim, Se Won Oh, Yoonho Nam, Hyun Gi Kim

**Affiliations:** Department of Radiology, Eunpyeong St. Mary’s Hospital, College of Medicine, The Catholic University of Korea, Seoul, Republic of Korea; Department of Biomedical Engineering, Hankuk University of Foreign Studies, Korea

**Keywords:** Hypoxic-ischemic injury, Glymphatic system, Perivascular space, Neonatal brain

## Abstract

**Objective:** To evaluate the relationship between hypoxic-ischemic injury (HII) and glymphatic function in neonates using two imaging biomarkers—basal ganglia perivascular space (BG-PVS) volume and diffusion-tensor imaging along the perivascular space (DTI-ALPS)—and to explore their interconnection.

**Methods:** This retrospective single-institution study collected neonatal brain MRIs from July 2020 to July 2022. BG-PVS volume and fraction were automatically extracted from 3D T2-weigthed images processing. DTI-ALPS indices were derived from DTI maps. Comparisons of BG-PVS parameters and DTI-ALPS indices were performed between neonates with and without HII. Logistic regression adjusted for age, sex, birth weight, and delivery mode. Correlations between BG-PVS parameters and DTI-ALPS indices were also evaluated.

**Results:** The study included 97 neonates without HII (median gestational age 252 days; 46 males) and 30 with HII (252 days; 16 males). Neonates with HII had smaller BG-PVS volumes (19 vs. 33 mm³, *p* = 0.001) and fractions (0.29% vs. 0.54%, *p* = 0.003) compared to neonates without HII. Logistic regression revealed a negative association between BG-PVS volume (OR: 0.96, 95% CI: 0.93–0.99) and fraction (OR: 0.15, 95% CI: 0.03–0.79) with HII. DTI-ALPS indices showed no evidence of differences between the neonates with and without HII (*p* = 0.54) or association with HII (*p* = 0.41). BG-PVS parameters and DTI-ALPS indices showed no evidence of correlation (coefficient = -0.28 – -0.08, *p*s > 0.05).

**Conclusion:** Neonates with HII demonstrated smaller BG-PVS volume and fraction compared to those without injury, indicating potential alterations in glymphatic function among affected newborns.

**Question:** Hypoxic-ischemic injury in neonates is a critical clinical concern, yet the relationship between this injury and glymphatic function remains unclear

**Findings:** Neonates with hypoxic-ischemic injury showed smaller basal ganglia perivascular spaces, while diffusion-tensor imaging analysis along perivascular space showed no significant difference.

**Clinical Relevance:** Basal ganglia perivascular space volume assessment provides a potential imaging biomarker for evaluating glymphatic system alterations in neonates with hypoxic-ischemic injury.

## Introduction

Neonatal brain injury can result from various processes, with hypoxia-ischemia being a significant cause. Neonatal encephalopathy, often caused by perinatal hypoxic-ischemic injury (HII) affect approximately 3 per 1,000 live births in high-income countries [1]. About 25% of neonates with hypoxia–ischemia encephalopathy exhibits subsequent neurological disabilities [2; 3], ranging from cognitive deficits to behavioral abnormalities. Additionally, children with a history of neonatal HII are more likely to develop attention-deficit/hyperactivity disorder and autism spectrum disorders [4]. Although long-term neurodevelopmental assessment is the gold standard for identifying impairments and disabilities, early detection and intervention offer a better chance to improve outcomes [5]. Therefore, a key area of research has focused on identifying potential imaging biomarkers through brain MRI.

The glymphatic system, a glial cell-dependent perivascular network responsible for removing brain waste products, is critical to neuronal health and has been linked to a variety of pathologies in children [6]. Its functionality is highly dependent on the structural integrity of the perivascular spaces (PVSs), which act as pathways for the circulation of fluids and the clearance of metabolic waste products [7]. Brain MRI has been used for studying the glymphatic system and its components, including MRI-visible PVS volume [8] and diffusion-tensor imaging (DTI) analysis along the perivascular space (DTI-ALPS) [9]. In the neonatal brain, the glymphatic system is thought to play an important role due to the high metabolic demands of rapid growth and development [10]. Studies showing visibility and volume of PVS [11] and DTI-ALPS indices [12] gave insights into glymphatic function change according to age and preterm birth. A recent study also showed a lower DTI-ALPS indices in neonates with birth asphyxia compared to those without [13]. However, since birth asphyxia does not always lead to HII evident on brain MRI and the study only included preterm neonates, we lack evidence to determine whether HII is associated with the glymphatic system in both preterm and term neonates. In addition, prior studies evaluated either PVS volume [8; 11] or the DTI-ALPS indices [12; 13], making it difficult to comprehensively assess the glymphatic system and evaluate the relationship between the two imaging biomarkers.

Therefore, the aim of this study was to investigate potential alternations in glymphatic system function among neonates with HII through analyzing two imaging biomarkers; basal ganglia perivascular space (BG-PVS) measurements and the DTI-ALPS. Specifically, we investigate the relationship between them as potential markers of glymphatic function.

## Materials and Methods

### Study Sample

The Institutional Review Board of our institution approved this retrospective study and informed consent was waived. This study was conducted in accordance with the Declaration of Helsinki. The study population consisted of neonates admitted to the Neonatal Intensive Care Unit (NICU) who underwent brain MRI between July 2020 and July 2022. All scans were performed for clinical purposes based on the neonatologist’s decision. The clinical indications included suspicion of brain injury, neurological symptoms, or abnormal findings on brain ultrasound. Initially, 254 neonates who underwent brain MRI during the study period were identified. We excluded 40 neonates who underwent brain MRI due to congenital abnormalities, follow-up studies, meningitis, seizures, metabolic disorders, and trauma. An additional 33 neonates were excluded as their brain MRIs were not obtained at term-equivalent age, and 14 more were excluded due to MRI motion artifacts or the absence of DTI data. Then neonates were categorized into those with and without MRI-visible HII. A flow chart for patient selection is shown in Fig 1.

**Figure 1.**
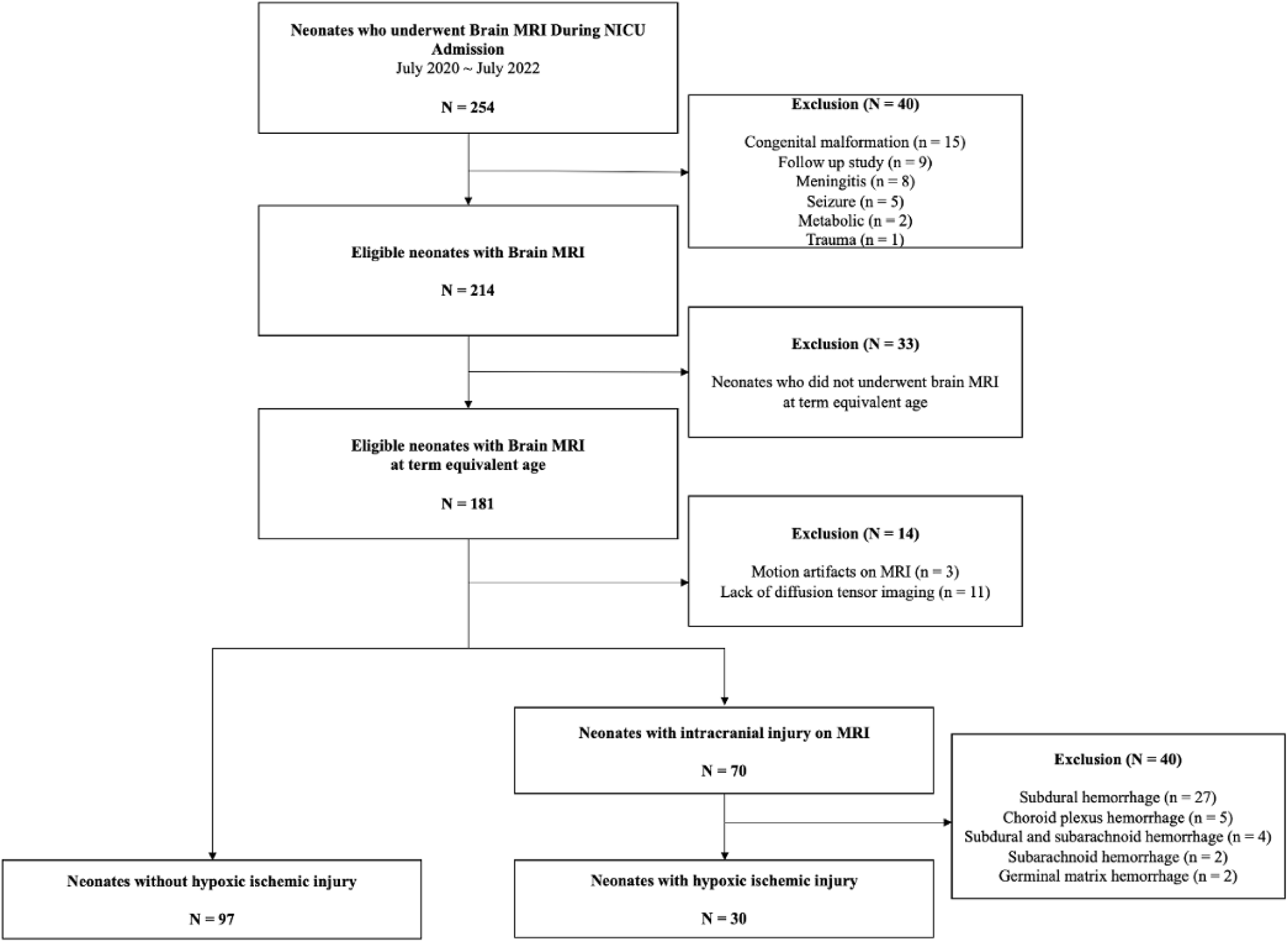
Flow chart of study group selection. NICU: neonatal intensive care unit

Neonates with MRI-visible HII were defined as those with the following brain MRI findings [14]: punctate white matter lesion, brain parenchyma cystic change, or intraventricular hemorrhage grade 3 or higher. Neonates with minor brain injuries with minor hemorrhages including small amount of subdural hemorrhage or choroid plexus hemorrhage were excluded. The final study sample included 127 neonates, with 97 neonates without HII and 30 neonates with HII.

Clinical information for the neonates, including sex, gestational age (GA) at birth, corrected gestational age (CGA) at scan, birth weight, mode of delivery, and Apgar scores (1 minute and 5 minutes), was collected.

### MRI acquisition

All MRI data were obtained using a 3T MRI scanner (Magnetom Vida, Siemens Healthineers). Neonates were scanned without sedation using a customized 64-channel receiver coil. 3D T1-weighted images (Magnetization-Prepared Rapid Acquisition with Gradient Echo, or MPRAGE), 3D T2-weighted images (Sampling Perfection with Application-Optimized Contrasts using Different Flip Angle Evolution, or SPACE), DTI, and susceptibility-weighted images (SWI) were obtained. Detailed imaging protocols are described in the Appendix.

### Automated Volumetric Measurements of PVSs

We first automatically segmented brain regions using the Infant FreeSurfer [15] and then Frangi filtering was performed to highlight enlarged perivascular space voxels [11; 16]. After Frangi filtering, we performed multiple thresholding by adding multiples of the standard deviation to the mean in the BG region (including caudate, putamen, and pallidum) to extract relatively high intensity voxels. Then, a researcher (*BLINDED*) performed manual refinement under the guidance of a radiologist with 15 years of experience in pediatric neuroradiology (*BLINDED*) to improve quantification accuracy. This process included removing false positive PVS voxels, correcting false negative PVS voxels, and determining the optimal thresholding values for enhancing label consistency of BG and PVS boundaries. The BG-PVS volume was calculated by counting the number of extracted voxels in the unit voxel volume. Values for PVS volume were collected from both hemispheres, and their sum values. The BG-PVS fraction was calculated by dividing the PVS volume by the segmented BG volume. Manual refinement was performed using ITK-SNAP version 4.2.0 [17], while other processes were performed using Python version 3.8 (Python Software Foundation).

### DTI Analysis along the Perivascular Space Index

The DTI-ALPS index assesses glymphatic system activity within the PVS by analyzing multidirectional diffusivity profiles generated from DTI data and we followed previous studies for DTI-ALPS index calculation [9; 12; 18]. SWI served as a reference for identifying direction of horizontal medullary vein for region of interest (ROI) placement. An axial section at this level was selected on the color-coded, DTI-derived fractional anisotropy (FA) map, aligning with the SWI section. On this axial image, two readers (*BLINDED READER 1* and *BLINDED READER 2*) independently placed three spherical ROIs (3–4 mm diameter) in the projection, association, and subcortical neural fiber regions of the left hemisphere. Using a custom software named DTI-ALPS Analyzer, based on MATLAB 9.6 (version 2019a, The Mathworks, Inc), nine distinct DTI-ALPS indices were automatically calculated from these ROIs: (a) Dx in the projection (Dxproj), association (Dxassoc), and subcortical (Dxsubc) neural fibers; (b) Dy in the same regions (Dyproj, Dyassoc, Dysubc); and (c) Dz in the same regions (Dzproj, Dzassoc, Dzsubc). The ALPS index was derived using the formula: ALPS index = mean (Dxproj, Dxassoc) / mean (Dyproj, Dzassoc). Reader 1 performed a second set of DTI-ALPS measurements one month after the initial assessment. The DTI-ALPS indices of reader 1 were used for analysis of association with HII and correlation with PVS parameters. The DTI-ALPS indices from reader 1’s first and second assessments were used to evaluate intraobserver agreement, and indices from reader 1 and reader 2 were analyzed to assess interobserver agreement.

### Statistics

Continuous variables were expressed as mean ± standard deviation (SD) or median [interquartile range (IQR)], and categorical variables as frequencies and percentages. Normality was assessed using the Shapiro-Wilk test. The sample size calculation was based on previously reported differences in PVS volume between preterm and term neonates [11], which indicated an effect size (Cohen’s d) of 0.47. With a sample size of 97 controls and 30 patients in the HII group (total N=127, α=0.05), we achieved 71.2 % power to detect similar effect sizes and were able to detect effect sizes of 0.67 or larger with 80 % power. This was considered adequate because HII may exert larger effects on PVS than GA difference. For group comparisons, we used Chi-square tests for categorical variables, t-tests for normally distributed continuous variables, and Mann-Whitney U tests for non-normal or ordinal variables. We performed a logistic regression analysis to evaluate the associations between HII in neonates and BG-PVS parameters along with DTI-ALPS. The main independent variables were BG-PVS volume, BG-PVS fraction, and DTI-ALPS indices and potential confounding factors including sex, GA, CGA, birth weight, and mode of delivery were adjusted. To quantify the relationships between each variable and the outcome, odds ratios (OR) with 95% confidence intervals (CI) were calculated, and statistical significance was assessed using p-value. Relationships between BG-PVS parameters and DTI-ALPS indices were assessed using Spearman’s correlation analysis, with results visualized using scatter plots and regression lines. Intra- and interobserver agreement of DTI-ALPS index was assessed using intraclass correlation coefficient (ICC) with a two-way random effects model for absolute agreement. ICC values were interpreted as: excellent (≥0.75), good (0.60-0.74), fair (0.40-0.59), and poor (<0.40), with 95% CI calculated for all measurements [19]. All tests were two-tailed, with significance at *p* < .05. Analyses were performed using R version 4.4.0 (R Core Team, 2024) by *BLINDED* with 10 years of statistical experience.

## Results

### Characteristics of Study Sample

Clinical and demographic characteristics are summarized in Table 1. There were 127 neonates (median GA [IQR], 252 [240;266] days, 62 males) in total, 97 neonates without HII (median GA [IQR], 252 [241;264] days, 46 males), and 30 neonates with HII (median GA [IQR], 252 [239;267] days, 16 males).

**Table 1.**
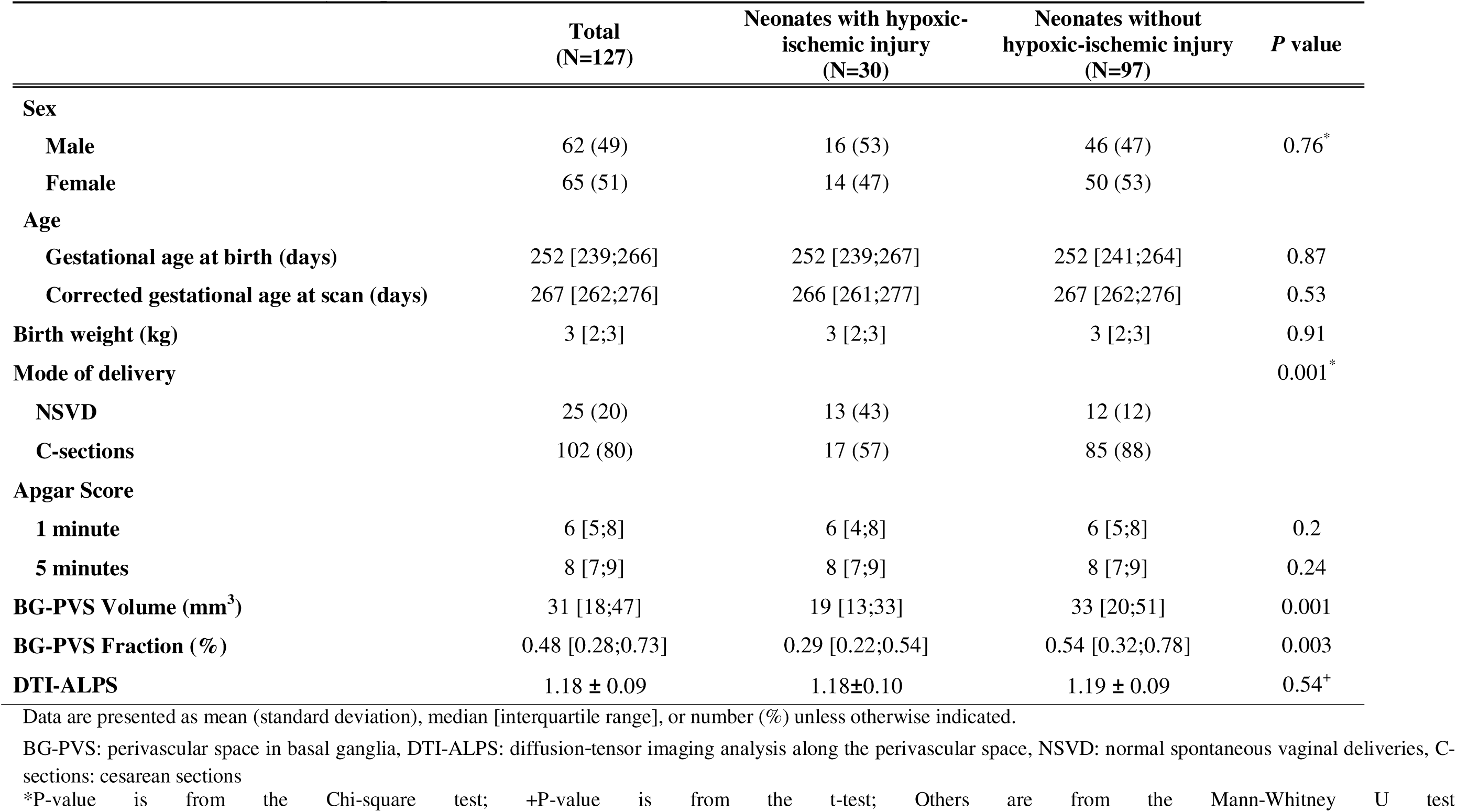
Characteristics of study sample.

|  | **Total**<br>**(N=127)** | **Neonates with hypoxic-**<br>**ischemic injury**<br>**(N=30)** | **Neonates without**<br>**hypoxic-ischemic injury**<br>**(N=97)** | **P value** |
| --- | --- | --- | --- | --- |
| **Sex** |  |  |  |  |
| **Male** | 62 (49) | 16 (53) | 46 (47) | 0.76* |
| **Female** | 65 (51) | 14 (47) | 50 (53) |  |
| **Age** |  |  |  |  |
| **Gestational age at birth (days)** | 252 [239;266] | 252 [239;267] | 252 [241;264] | 0.87 |
| **Corrected gestational age at scan (days)** | 267 [262;276] | 266 [261;277] | 267 [262;276] | 0.53 |
| **Birth weight (kg)** | 3 [2;3] | 3 [2;3] | 3 [2;3] | 0.91 |
| **Mode of delivery** |  |  |  | 0.001* |
| **NSVD** | 25 (20) | 13 (43) | 12 (12) |  |
| **C-sections** | 102 (80) | 17 (57) | 85 (88) |  |
| **Apgar Score** |  |  |  |  |
| **1 minute** | 6 [5;8] | 6 [4;8] | 6 [5;8] | 0.2 |
| **5 minutes** | 8 [7;9] | 8 [7;9] | 8 [7;9] | 0.24 |
| **BG-PVS Volume (mm<sup>3</sup>)** | 31 [18;47] | 19 [13;33] | 33 [20;51] | 0.001 |
| **BG-PVS Fraction (%)** | 0.48 [0.28;0.73] | 0.29 [0.22;0.54] | 0.54 [0.32;0.78] | 0.003 |
| **DTI-ALPS** | 1.18 ± 0.09 | 1.18±0.10 | 1.19 ± 0.09 | 0.54<sup>+</sup> |

Data are presented as mean (standard deviation), median [interquartile range], or number (%) unless otherwise indicated.
BG-PVS: perivascular space in basal ganglia, DTI-ALPS: diffusion-tensor imaging analysis along the perivascular space, NSVD: normal spontaneous vaginal deliveries, C-sections: cesarean sections
\*P-value is from the Chi-square test; +P-value is from the t-test; Others are from the Mann-Whitney U test

Gestational age, CGA, birth weight, and 1 minute and 5 minutes Apgar scores did not show evidence of difference between neonates with and without HII (all *p*s > 0.05) (Table 1). Birth type was different between the two groups with and without HII showing more neonates born cesarean sections in neonates without HII (neonates with HII vs. neonates without HII; 17 [57 %] vs. 85 [88 %], *p =* 0.001).

### Difference in Glymphatic Parameters Between Neonates With and Without Hypoxic-Ischemic Injury

When comparing glymphatic parameters, the BG-PVS volume and fraction differed between neonates with and without HII, while the DTI-ALPS indices showed no evidence of difference (Table 1 and Fig 2). The neonates with HII had smaller BG-PVS volume (neonates with HII vs. neonates without HII; median [IQR], 19 [13;33] mm³ vs. 33 [20;51] mm³, *p =* 0.001) and BG-PVS fraction (neonates with HII vs. neonates without HII; median [IQR], 0.29 [0.22;0.54] % vs. 0.54 [0.32;0.78] %, *p =* 0.003) compared to the neonates without HII. DTI-ALPS indices did not show evidence of difference between the two groups (neonates with HII vs. neonates without HII; mean ± SD, 1.18±0.10 vs. 1.19±0.09, *p =* 0.54). Representative images of a neonate with HII and a neonate without HII are shown in Fig 3 and Fig 4, respectively.

**Figure 2.**
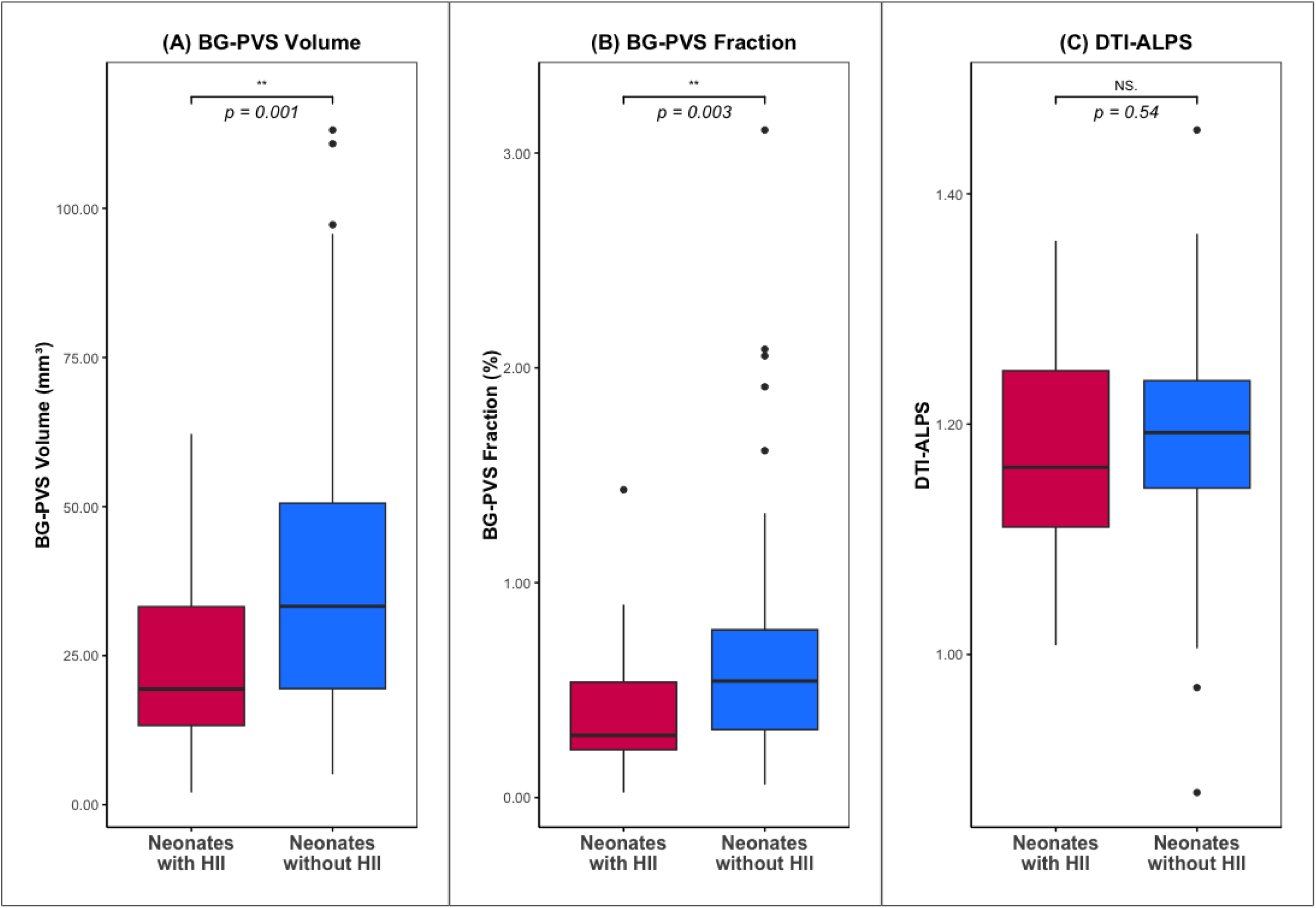
Box plots comparing glymphatic parameters between neonates with and without hypoxic-ischemic injury. There were differences in BG-PVS volume (A) and BG-PVS fraction (B) between neonates with and without HII. However, the DTI-ALPS (C) showed no evidence of difference between the two groups. BG-PVS: perivascular space in basal ganglia; DTI-ALPS: diffusion-tensor imaging analysis along the perivascular space; HII: hypoxic-ischemic injury.

**Figure 3.**
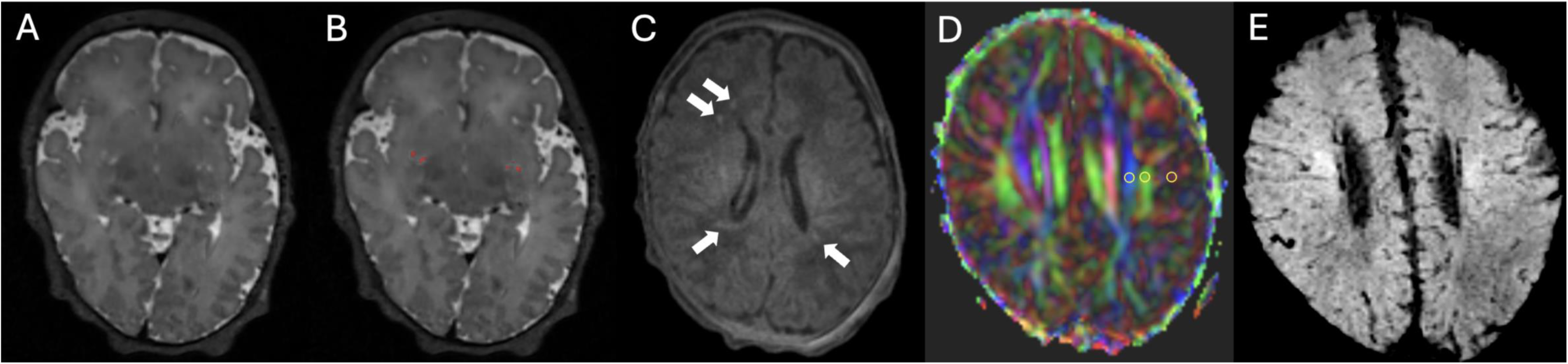
Representative brain MRI images of a female neonate with hypoxic ischemic injury (born at 38 weeks GA, MRI performed at 41 weeks CGA). An axial T2-weighted image shows BG-PVS (A), and the areas are segmented in red (B). A T1-weighted image (C) shows hyperintense lesions in both periventricular white matter (arrows), suggesting hypoxic ischemic injury. A color-coded, DTI-derived fractional anisotropy map with ROIs (yellow circles) for DTI-ALPS metrics is shown (D), accompanied by a reference SWI image illustrating medullary vein direction (E). The three ROIs represent projection, association, and subcortical fibers from left to right. Diffusivity of the projection and association fibers was used to calculate the DTI-ALPS index. The segmented BG-PVS was 25 mm^3^ with a fraction of 0.3 %. DTI-ALPS index was 1.14. BG-PVS: perivascular space in basal ganglia; CGA: corrected gestational age; DTI-ALPS: diffusion-tensor imaging analysis along the perivascular space; GA: gestational Age; ROI: region of interest.

**Figure 4.**
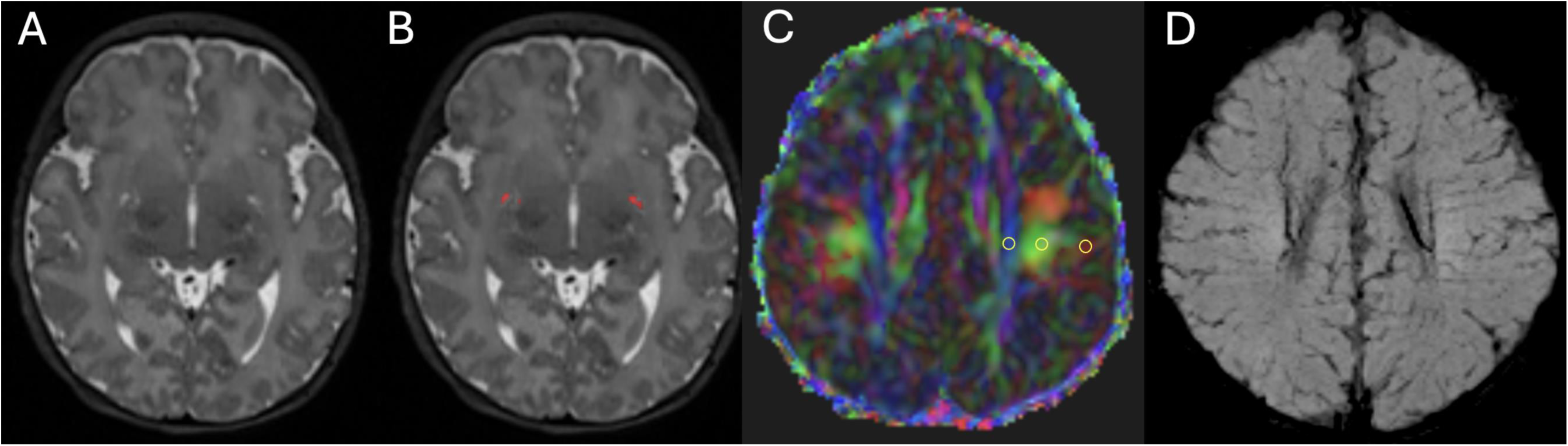
Representative brain MRI images of a female neonate without hypoxic ischemic injury (born at 39 weeks GA, MRI performed at 41 weeks CGA). An axial T2-weighted image shows BG-PVS (A) and the areas are segmented in red (B). A color-coded, DTI-derived fractional anisotropy map with ROIs (yellow circles) for DTI-ALPS metrics is shown (C) with a reference SWI image showing medullar vein direction (D). The segmented BG-PVS measured 39 mm^3^ with a fraction of 0.7 %. DTI-ALPS index was 1.17. The three ROIs represent projection, association, and subcortical fibers from left to right. Diffusivity of the projection and association fibers was used to calculate the DTI-ALPS index. BG-PVS: perivascular space in basal ganglia; CGA: corrected gestational age; DTI-ALPS: diffusion-tensor imaging analysis along the perivascular space; GA: gestational Age; ROI: region of interest.

### Associations between Glymphatic Parameters and Hypoxia-Ischemic Injury

Logistic regression analysis revealed a negative association between BG-PVS volume and HII (OR: 0.96, 95% CI: 0.93−0.99, *p* = 0.007) (Table 2). Similarly, BG-PVS fraction and HII showed a negative association (OR: 0.15, 95% CI: 0.03−0.79, *p =* 0.026). DTI-ALPS indices and HII did not show evidence of association (OR: 0.10, 95% CI: 0.00−25.41, *p =* 0.411).

**Table 2.** Associations between glymphatic parameters and hypoxic-ischemic injury in neonates.

| Variable | OR (95% CI) | *P* value |
| --- | --- | --- |
| **BG-PVS Volume** | 0.96 (0.93–0.99) | 0.007 |
| **BG-PVS Fraction** | 0.15 (0.03–0.79) | 0.026 |
| **DTI-ALPS** | 0.10 (0.00–25.41) | 0.411 |

OR: odds ratio, CI: Confidence Interval, BG-PVS = perivascular space in basal ganglia, DTI-ALPS: diffusion-tensor imaging analysis along the perivascular space
All models are adjusted for sex, corrected gestational age, birth weight, and mode of delivery
BG-PVS Volume is measured in mm<sup>3</sup> and BG-PVS Fraction is expressed as a percentage.

### Correlations between Glymphatic Parameters

The relationships between ‘BG-PVS volume and DTI-ALPS indices’ and ‘BG-PVS fraction and DTI-ALPS indices’ are shown in Fig 5. BG-PVS volume and DTI-ALPS indices did not show evidence of correlation in neonates in total, neonates without HII, or neonates with HII (range of r = -0.28 − -0.08, *p*s > .05). Similarly, the BG-PVS fraction did not show evidence of correlations with DTI-ALPS indices in all three groups of neonates (range of r = -0.18 − -0.15, *p*s > .05).

**Figure 5.**
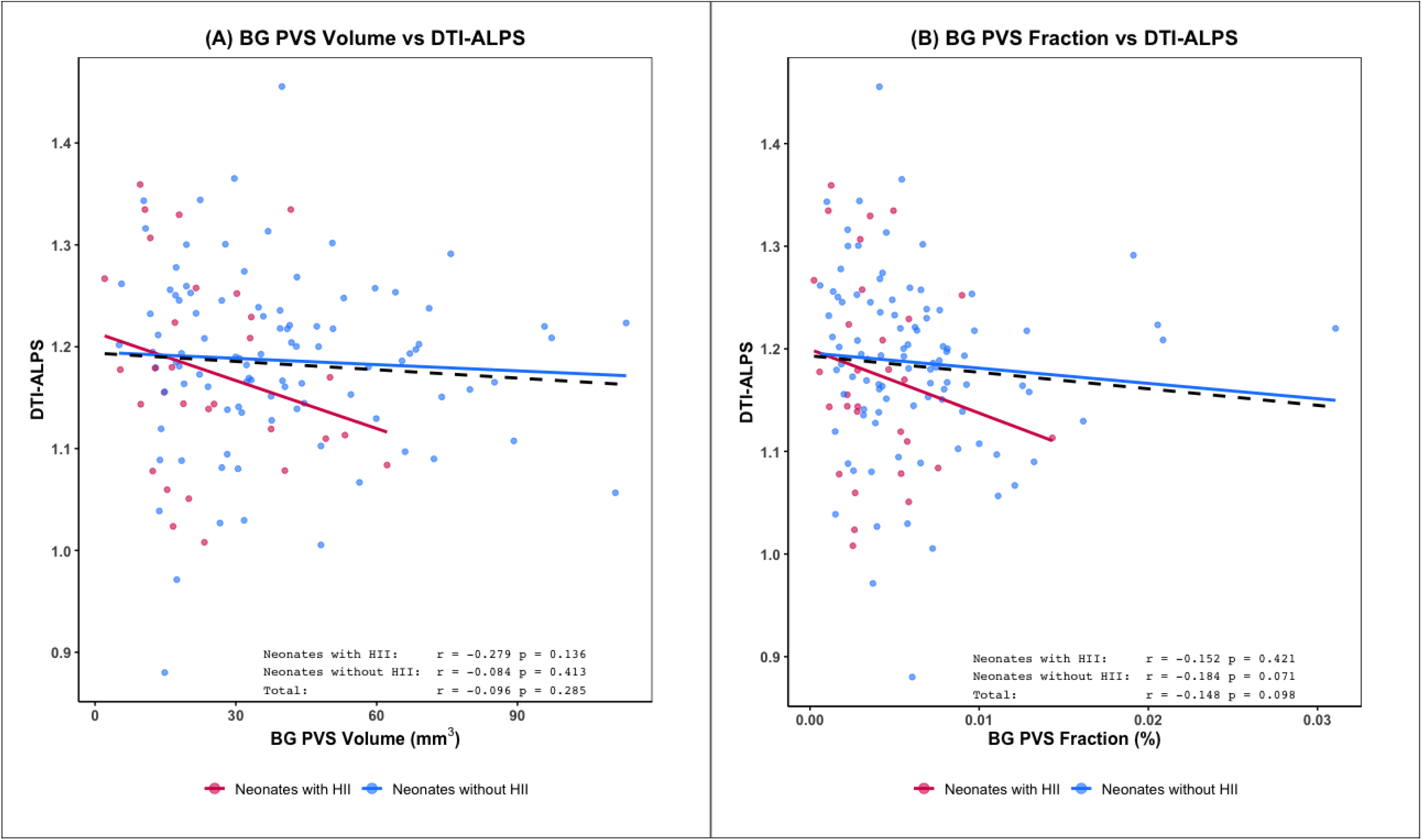
Scatter plots illustrating the relationship between BG-PVS parameters and the DTI-ALPS index. No significant correlations were observed between BG-PVS volume and the DTI-ALPS index (A) or between BG-PVS fraction and the DTI-ALPS index (B). Red dots and trend lines represent neonates with HII, while blue dots and trend lines represent neonates without HII. The dashed black lines indicate the overall trend for all neonates. BG-PVS: perivascular space in basal ganglia; DTI-ALPS: diffusion-tensor imaging analysis along the perivascular space; HII: hypoxic-ischemic injury.

### Intra- and Interobserver agreement of DTI-ALPS index

Intra- and interobserver agreement analysis demonstrated excellent reliability for DTI-ALPS indices. The intraobserver agreement showed excellent reliability with an ICC of 0.825 (95% CI: 0.76−0.87). The interobserver agreement also showed excellent reliability with an ICC of 0.868 (95% CI: 0.82−0.91).

## Discussion

Brain glymphatic function may be altered following hypoxic-ischemic injury (HII) in neonates. However, imaging biomarkers such as basal ganglia perivascular space (BG-PVS) volume and diffusion-tensor imaging analysis along the perivascular space (DTI-ALPS) have not been studied for their association with HII. Additionally, the relationship between these two imaging markers has not been investigated. We found that neonates with HII had smaller lower BG-PVS volume (*p* = 0.001) and fraction (*p* = 0.003) compared to those without HII. Logistic regression analysis revealed a negative association between BG-PVS volume (OR: 0.96, 95% CI: 0.93–0.99) and fraction (OR: 0.15, 95% CI: 0.03–0.79) with HII, while DTI-ALPS indices showed no evidence of association (OR: 0.10, 95% CI: 0.00–25.41, *p* = 0.411). We found no evidence of correlations between BG-PVS parameters and DTI-ALPS indices (coefficient = -0.28 − -0.08, *p*s > 0.05).

Our study revealed significant differences in glymphatic system parameters of BG-PVS volume and fraction between neonates with and without HII indicating that HII may lead to alterations in the glymphatic system. This result was similar to a previous study showing that neonates with preterm birth have smaller BG-PVS volumes compared to controls [11]. The finding of smaller BG-PVS volume in diseased neonates contrasts with findings in older children, who exhibit larger PVS volumes in disorders such as attention-deficit/hyperactivity disorder [20] or autism [21]. Therefore, we can assume that altered glymphatic function may affect PVS volume but will have different affect depending on the age of a subject. In neonatal brain, smaller BG-PVS volume can be attributed to abnormally high expression of aquaporin-4 which mediate influx of cerebrospinal fluid into PVS. In addition, ischemia injury can lead to astrocyte swelling [22] which may result in dysfunction of or immature glymphatic system.

While PVS parameters revealed differences between the two groups of neonates with and without HII, no such differences were observed in DTI-ALPS indices. This finding contrasts with a previous study that demonstrated a lower DTI-ALPS indices in neonates with birth asphyxia compared to those without (neonates with asphyxia vs. without asphyxia; 0.98 ±D0.08 vs. 1.08 ±D0.07, *p* < .05) [13]. This discrepancy can be attributed to several factors including variations in subject characteristics and MRI timing. Unlike the previous study, which involved preterm neonates with a median GA of 29-30 weeks, our neonates had a median GA of 36 weeks. Additionally, MRIs in the previous study were acquired at a median age of 31 weeks, whereas our neonates were scanned at term-equivalent age. Based on our study results, the BG-PVS volume or fraction values appears to be a more sensitive marker for detecting glymphatic alterations in neonates who undergo brain MRI at term-equivalent age. However, further studies evaluating both BG-PVS and DTI-ALPS indices across a range of GA and CGA at the time of MRI are needed to confirm these findings.

Our logistic regression analysis revealed association between BG-PVS parameters and HII in neonates after adjusting sex, GA, CGA, birth weight, and mode of delivery. The initial group comparison showed no demographic differences between the two neonatal groups (with and without HII), except for mode of delivery. The logistic regression analysis, adjusted for potential confounding variables, further clarified the association between BG-PVS parameters and HII, suggesting BG-PVS parameters may serve as independent imaging markers associated with HII. To establish the predictive value of neonatal glymphatic function on HII, a longitudinal study with early glymphatic function parameter measurements and subsequent outcome assessments will be needed. Additionally, since HII may result in neurodevelopmental disorders [2; 3], it is important to explore whether BG-PVS parameter values impact the likelihood of such disorders following HII.

Given that both BG-PVS and the DTI-ALPS indices have shown promising findings in neonates, with age associations reported in two separate studies [11; 12], we assumed a relationship between BG-PVS and the DTI-ALPS indices. However, we found no evidence of correlation between BG-PVS parameters and DTI-ALPS indices. A previous study of adults with Parkinson’s disease, which showed that a decrease in the DTI-ALPS indices correlated with an increased PVS burden [23]. Our findings suggest that the relationship between BG-PVS and the DTI-ALPS indices may differ in neonates, partly due to the maturation process of glymphatic pathways and perivascular spaces over time. Consequently, their interaction in the neonatal period may not reflect patterns observed in older populations. In addition, as PVS in different brain regions (BG vs. white matter) is associated with different diseases [24], and since BG-PVS (measured in BG) and DTI-ALPS (measured in white matter) reflect the glymphatic system in distinct areas, our finding of no correlation between the two may suggest differing roles and conditions in these regions in neonates. Due to the limited visibility of white matter PVS on neonatal brain MRI [11] quantifying PVS volume in this region and evaluating its correlation with the DTI-ALPS index is currently challenging. Larger-scale longitudinal studies would be needed to clarify the relationship between different glymphatic system markers in neonates and to provide a comprehensive understanding.

Our study had several limitations. First, as a cross-sectional retrospective study, it was difficult to establish a causal relationship between MRI-derived glymphatic markers and HII in neonates. Although smaller BG-PVS parameters may result from HII, a clear relationship should be explored through studies assessing glymphatic parameters earlier in the perinatal period. Second, PVS was measured only in the BG and not in white matter as discussed above. Consequently, whether white matter PVS plays a distinct role, undergoes different alterations in glymphatic function in neonates, or has a relationship with DTI-ALPS indices remains unclear.

In conclusion, hypoxic ischemic injury (HII) in neonates was associated with smaller basal ganglia perivascular space (BG-PVS) volumes and fractions. The diffusion-tensor imaging analysis along the perivascular space (DTI-ALPS) indices showed no association with HII, and no correlation was observed between BG-PVS parameters and the DTI-ALPS indices. These findings suggest that glymphatic function may be altered in association with HII in neonates. Our results also indicate distinct roles for BG-PVS and DTI-ALPS in neonatal brain pathology, implying they may reflect different aspects of glymphatic function. Future research with longitudinal studies is warranted to explore how these markers evolve with maturation and their potential predictive value for neonatal neurodevelopmental outcome.

## Data Availability

All data produced in the present study are available upon reasonable request to the authors

## Abbreviations

*BG-PVS*: Basal ganglia perivascular space
*CGA*: Corrected gestational age
*CI*: Confidence intervals
*DTI-ALPS*: Diffusion-tensor imaging analysis along the perivascular space;
*GA*: Gestational age
*HII*: Hypoxic-ischemic injury
*IQR*: Interquartile range
*NICU*: Neonatal intensive care unit
*NSVD*: Normal spontaneous vaginal delivery
*OR*: Odd ratio
*SD*: Standard deviation.

